# Epidemiology and risk factors for diarrhoeagenic *Escherichia coli* carriage among children in northern Ibadan, Nigeria

**DOI:** 10.1101/2022.09.26.22280249

**Authors:** Olabisi C. Akinlabi, El-shama Q. Nwoko, Rotimi A. Dada, Stella Ekpo, Adeola Omotuyi, Akinlolu Adepoju, Oluwafemi Popoola, Gordon Dougan, Nicholas R. Thomson, Iruka N. Okeke

## Abstract

Diarrhea is a leading cause of childhood morbidity in Africa. Outside of multi-country sentinels, of which there are none in Nigeria, few studies focus on bacterial etiology. We performed a case-control study among children under five years of age. Stool specimens were collected from 120 children with, and 357 without, diarrhea attending primary health clinics on the northern outskirts of Ibadan between November 2015 and August 2019. Up to ten *E. coli* isolates were obtained per specimen and at least three were whole genome-sequenced using Illumina technology. Genomes were assembled using SPAdes, quality evaluated using QUAST, and Virulencefinder was used to identify virulence genes. The microbiological quality of water from 14 wells within the study area was assessed using total and coliform counts. Diarrhoeagenic *Escherichia coli* (DEC) were isolated from 79 (65.8%) of cases and 217 (60.8%) control children. All DEC pathotypes except Shiga toxin-producing *E. coli*, a number of hybrid DEC pathotypes, *Salmonella* and *Yersina* spp. were detected but no pathogen showed association with disease (p>0.05). Enterotoxigenic *E. coli* were more commonly recovered from younger controls but exclusively detected in cases aged over nine months. Temporally-linked, highly similar enteroaggregative *E. coli* were isolated from children in different households in eight instances. No well water sample drawn in the study qualified as potable. Children in northern Ibadan are commonly colonized with DEC. Access to water and sanitation, and vaccines targeting the most abundant pathogens may be critical for protecting children from the less overt consequences of enteric pathogen carriage.

## Introduction

Diarrhea has been reported to kill about 480,000 children each year worldwide and it accounted for 8% of all deaths in children less than five years of age in 2017.^1^ This translates to about 1,300 pediatric deaths each day, mainly in low- and middle-income countries, which are home to about 62% of the world’s population under five years of age. Nigeria is reported to have the greatest number of under-five deaths annually^2^, and in the year 2019 UNICEF attributed about 18% of this mortality to diarrhea.^3^ Global mortality from diarrhea declined by 61% between 1990 to 2020^2^ owing to improvements in preventive and treatment interventions. Improvements continue but they are seen less with morbidity in Nigeria. ^4,5^

Understanding the etiology of diarrhea can help to discern and interrupt transmission pathways, overcome pathogen-specific risk factors and provide valuable information to support the development of vaccines and therapeutics. The range of pathogens that can produce diarrhea is extensive and includes enterovirulent bacteria such as *Salmonella* and diarrhoeagenic *Escherichia coli*. Diarrhoeagenic *E. coli* (DEC) are classified, based on virulence gene content and/or epithileal cell adherence pattern, into different pathotypes including *Shigella* and enteroinvasive *E. coli* (EIEC), Shiga toxin-producing *E. coli* (STEC), enterotoxigenic *E. coli* (ETEC), enterohaemorrhagic *E. coli* (EHEC), enteropathogenic *E. coli* (EPEC) and enteroaggregative *E. coli* (EAEC). ^5^ The epidemiology of different DEC pathotypes varies geographically and few studies have attempted to identify risk factors for DEC in childhood diarrhea.^7-13^

DEC are particularly neglected diarrheal pathogens, in part because they are challenging to differentiate from commensal *E. coli*. There have only been a handful of reports from Nigeria on the role of DEC in diarrhea. They range from the earliest studies of Agbonlahor and Odugbemi (1982),^14^ Antai and Anozie (1987)^15^ and Agbodaze et al (1988)^16^ to work done in the last three decades by Ogunsanya et al (1994),^17^ Okeke et al (2000),^18^ Nweze (2010),^19^ Onanuga *et al* (2014),^20^ Ifeanyi et al (2015),^21^ and Odetoyin et al. (2016).^22^ Nigeria (including Ibadan) is reported to have a high burden of faeco-orally transmitted disease. ^23^ But studies have focused on only a subset of DEC pathotypes, collectively have studied a very limited number of locations and no diarrhea etiologic study has been performed in or around Ibadan.^24^ Additionally, most of the aforementioned studies have used methods that are insufficiently sensitive to reliably and comprehensively identify DEC pathotypes. HEp-2 cell adherence tests, Gold Standards for EPEC, EAEC and DAEC (but unable to identify other categories), are not performed in Nigeria and have only been used in one instance.^17^ The earliest studies used serotyping,^5,14,15,17,^ which is only partially predictive for a few pathogenic subtypes (notably typical EPEC), and while more recent studies did seek molecular targets, they based identification of DEC pathotypes on only one or two of such targets.^18,22,25-30^ This is sufficient for some pathotypes, such as EPEC, ETEC and EIEC, but is notably insensitive for EAEC, which available data suggest may be of significant epidemiological importance in this region, ^31^ and a neglected pathogen globally.^32^ This study aimed to look for associations of DEC pathotypes with diarrhea in children from Ibadan southwestern Nigeria as well as to also determine the social demographic risk factors associated with each DEC pathotypes. While we did not have access to HEp-2 adherence testing, we aimed to expand sensitivity by screening for DEC targets through whole genome sequencing (WGS).

## Materials and methods

### Study design and ethical approval

Ethical approval for this research was obtained from the University of Ibadan/ University College Hospital (UI/UCH) ethics committee (approval number UI/EC/15/093). This study was a case control study enrolling children up to 5 years old with and without diarrhea. Case faecal samples were collected from children between the age of 0-5 years, whose parents or guardians consented, who were diagnosed with acute diarrhea by a health worker or community health officer. Controls were children under five visiting the health center for vaccination or minor trauma and deemed otherwise healthy by well-baby practitioner. Children that were potentially eligible as cases or controls were excluded if they had been treated with antibiotics in the last one month. A detailed questionnaire was offered to obtain information on clinical history and physical examination findings.

### Sample sites and size

The study was conducted on the northern outskirts of Ibadan, the state capital of Oyo state commonly known as the city of the red roofs due to its ancient buildings. Five different primary health centers in Lagelu and Egbeda local government areas (LGAs), which lie immediately north east of metropolis (Figure 1) served as enrollment points for the study. These LGAs are semi urban and lack good access to water and sanitation. A total of 477 samples were collected from enrollees that comprise 120 case and 357 controls.

**Figure 1:**
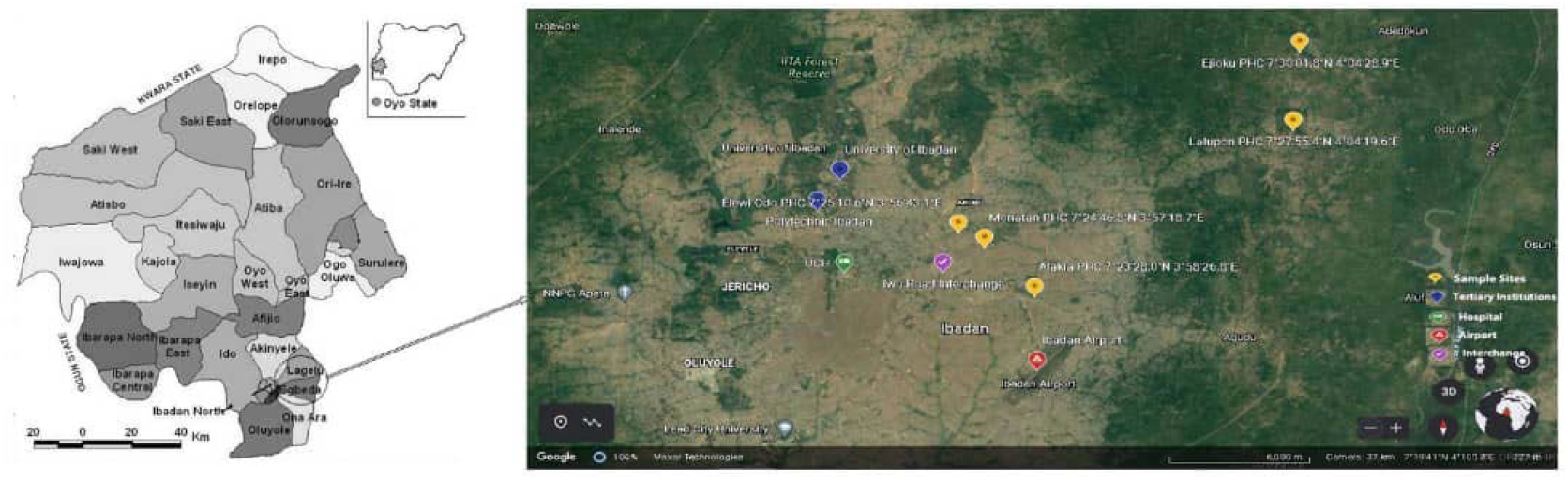
Study location (A) Ibadan metropolis is comprised of five central and six outlying local government areas. Lagelu and Egbeda are in the northeastern suburbs. (B) Map of the study area showing the location of the five participating primary health care centers.

### Sample processing, culture and bacterial isolation

Stool specimens were transported to the laboratory within 1-2 hours of collection, they were plated onto eosin methylene blue agar (Oxoid), MacConkey agar plates (Oxoid), Selenite F broth and incubated at 37 °C for 18 hours. Five representative non-lactose and five lactose fermenting colonies, capturing all morphological appearances (colour, shape, edge and colony surface) were picked and sub-cultured until pure cultures were obtained. Growth from the Selenite F broth was further streaked on Xylose Lysine Deoxycholate agar. Black colonies that were center dotted were obtained and isolate stocks were stored by freezing at -80 °C in Luria broth:glycerol 1:1. Species were identified biochemically using the Microbact 24E system (Oxoid) and *Salmonella* spp. were confirmed by *invA* PCR.^33^

### Occult blood testing

Occult blood was detected using a qualitative immunoassay kit for hemoglobin (Cromatest) in accordance with manufacturer instructions. Fresh stool samples were emulsified and placed on the sample section containing the particle coated with anti-hemoglobin antibody of the kit and a drop of the buffer was added to it then timed for one minute for the stool and buffer mix to migrate upward by capillary action. The presence of two lines (control and positive lines) indicates that the reaction is positive and a single line (control line) indicates a negative result.

### DNA extraction, sequencing and sequence analyses

All representative *E. coli* types discernable from colony morphology and Microbact 24 biochemical profiles from each specimen were sequenced. Where there were less than three different profiles, at least three *E. coli* isolates were sequenced per specimen. DNA extraction of *E. coli* isolates was done aseptically using the Wizard Genomic Extraction kit (Promega) according to manufacturers’ protocol and DNA samples were library prepared and whole genome sequenced using Illumina platform. Raw read quality control was carried out using FastQC (Babraham Bioinformatics, Babraham Institute, Cambridge, United Kingdom) and quality reports aggregated using MultiQC.^34^ Reads were assembled using SPAdes assembler and assembly quality was determined using Quality Assessment Tool for Genome Assemblies (QUAST),^35^ and CheckM.^36^ Reads were also assigned taxonomic Id using Kraken and the taxonomic ids assigned by Kraken,^37^ were used to determine species abundance using Bracken.^38^ ARIBA Virulencefinder database,^39^ was used to identify virulence genes. Isolate genomes were deposited in the European Nucleotide Archive (ENA) under the project ID PRJEB8667 (https://www.ebi.ac.uk/ena/browser/view/PRJEB8667).

### Identification of DEC pathotypes from whole genome sequence

The VirulencFinder output was used to classify *E. coli* into pathotypes. Sequenced *E. coli* isolates with any of the genes *aaiC, aar, aap, aatA (CVD432), aggA, aafA, agg3A, agg4A, agg5A, aggR, air, capU*, and *eilA* were classified as EAEC. *E. coli* with either Locus of Enterocyte Effacement (LEE) genes (including *eae*) and *bfp*, or LEE genes without *bfp* or *stx* genes are classified as EPEC. Those with either *sta* (enterotoxins heat stable (ST) of ETEC) and/or *ltcA* (enterotoxins heat labile (LT) of ETEC) with or without *lngA* were classified as ETEC. Sequenced *E. coli*/ *Shigella* with either *ipaD, ipaH* and/or *virF*, were classified as EIEC/*Shigella*. The presence of any *stx* gene would have categorized a strain as STEC, with those also harboring LEE genes considered EHEC.

### Sentinel water quality analysis

Samples from 14 water sources (all wells) that were proximal to the health centers were collected for analysis during the second year of the study. One liter of each water sample was collected aseptically into sterile bottles and transported to the laboratory on ice for analysis within an hour. pH was measured using the HI 2210 pH meter (Hanna instrument), total counts were computed after plating on tryptic soy agar (Oxoid) and coliform counts were performed on MacConkey agar plates (Oxoid). Isolates were identified as for stool isolates.

### Statistical Analysis

Epi Info version 7 software (Centers for Disease Control and Prevention, Atlanta, GA, USA) and SPSS (Statistical Package for Social Science) version 20 software were used for statistical calculations. Fisher exact test and Chi square test were used to test the association between the cases and control with various pathotypes and genes while bivalent analysis and logistic regression was used to analyze the significance of potential risk factors. P-values < 0.05 were considered to be statistically significant applying Bonferonni corrections, where appropriate.^40^

## RESULTS

### High rates of diarrhoeagenic *E. coli* recovery from cases and controls

Stool specimens from 120 consented patients with diarrhea and 357 apparently healthy controls attending primary health care facilities in Egbeda and Lagelu local government areas of Ibadan, Nigeria were processed between November, 2016 and August, 2019. Figure 2 shows how under-five case enrollment was distributed over the year from November 2015 to August 2019. As the figure indicates, there was a health system strike between 7^th^ of June 2016 and 25^th^ July 2016 during which time patients could not be recruited (Figure 2). The figure also shows temperature and rainfall distribution of Oyo state, over the sample collection period, revealing no association between rainfall, temperature and diarrhea cases recruitment during the study.

**Figure 2:**
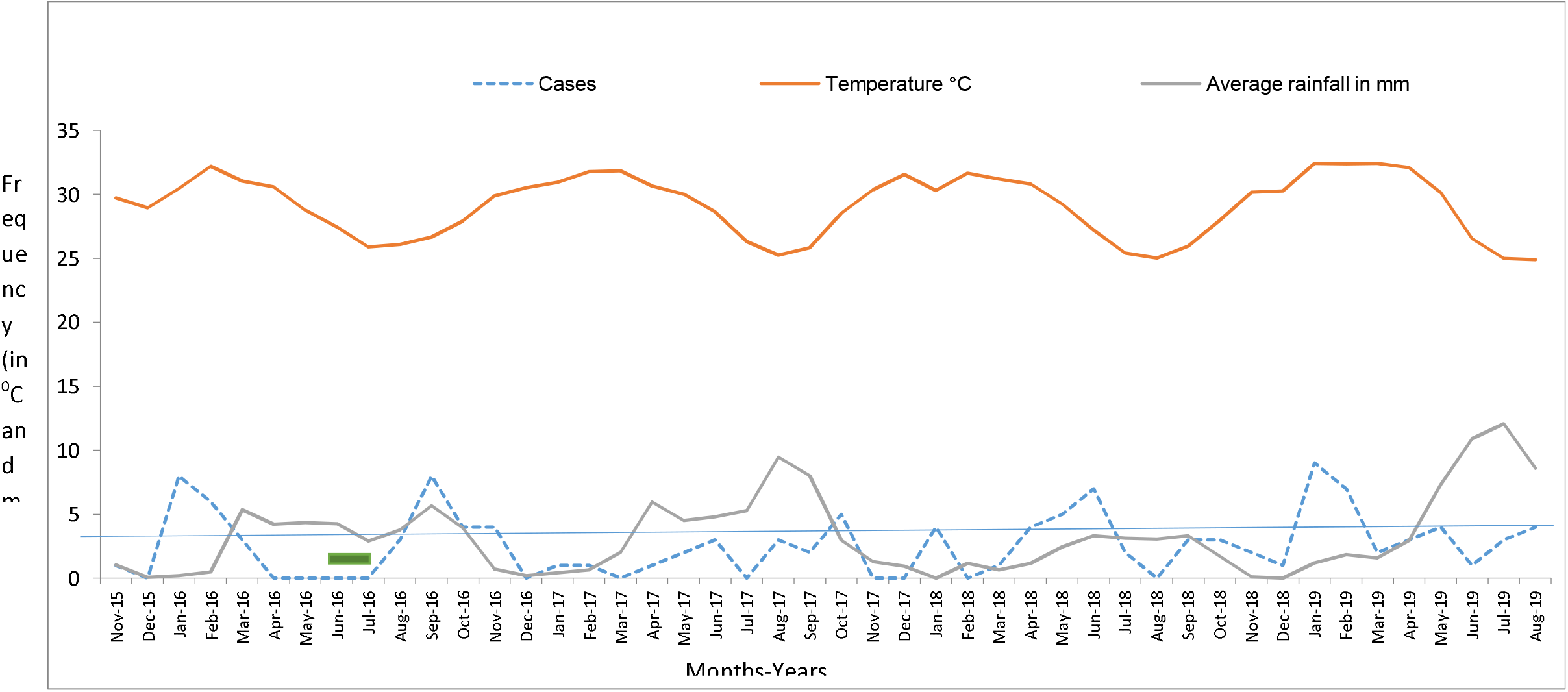
Number of cases collected per months over the course of the study including the average rainfall and temperature (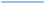 median).

DEC were identified from 323 (68.6%) of the stool specimens, and include EAEC 281 (58.9%), EPEC 23 (4.8%), ETEC 19 (4.0%) and EIEC 1 (0.2%). None of the sequenced *E. coli* carried the *stx1* and/or *stx2* genes (encoding Shiga toxins 1 and 2), which would have classified strains as STEC/EHEC. Table 1 shows the recovery of each diarrhoeagenic *E. coli* pathotype from cases and controls. EAEC was the most frequently recovered pathotype. ETEC strains were recovered from 8 cases and 11 controls (p= 0.0823) and ETEC bearing the ST-encoding gene (*sta*) were only isolated from patients with diarrhea. However, no pathotype was significantly associated with disease. A total of 32 isolates met the definition of more than one pathotype and would therefore be considered pathotype hybrids. With the exception of one ETEC-EPEC hybrid, the hybrids all carried EAEC genes, with EAEC-ETEC and EAEC-EPEC being the most common (Table I). No hybrid pathotype was associated with diarrhea.

**Table 1:** The DEC pathotypes isolated from the cases and control samples.

| Pathotypes | Cases (%)<br>[n=120] | Control (%)<br>[n=357] | Total<br>(477) | P-value | Odds<br>ratio | CI(Lower-<br>upper) |
| --- | --- | --- | --- | --- | --- | --- |
| EAEC | 72 (60) | 209 (58.5) | 281 | 0.7790 | 1.0622 | 0.6968-1.6192 |
| EAEC-EIEC | 0 | 1 | 1 | 1.0000 |  |  |
| EAEC-ETEC | 5 | 7 | 12 | 0.1876 |  |  |
| EAEC-EPEC | 4 | 6 | 10 | 0.2794 |  |  |
| EPEC | 7 (5.8) | 16 (4.5) | 23 | 0.5499 | 1.3202 | 0.5297-3.2908 |
| *eae* | 7 (5.8) | 16 (4.5) | 23 | 0.5499 |  |  |
| *bfp*+*eae*(Typical<br>EPEC) | 1 (0.8) | 2 (0.6) | 3 | 1 |  |  |
| ETEC | 8 (6.7) | 11 (3.1) | 19 | 0.0823 | 2.2468 | 0.8818-5.7248 |
| *sta* | 2 (1.7 ) | 0 (0) | 2 | 0.0629 |  |  |
| *ltcA* | 7 (5.8) | 11 (3.1) | 18 | 0.1711 |  |  |
| *sta*+*ltcA* | 1 (0.8) | 0 (0) | 1 | 0.2516 |  |  |
| *ltcA*+*lngA* | 1 (0.8) | 0 (0) | 1 | 0.2516 |  |  |
| ETEC-EPEC | 1 | 0 | 1 | 0.2526 |  |  |
| EIEC/ Shigella | 0 (0) | 1 (0.3) | 1 | 0.3368 | 0.0000 | Inf-inf |
| *ipaH* | 0 (0) | 1 (0.3) | 1 |  |  |  |
| *ipaD* | 0 (0) | 1 (0.3) | 1 |  |  |  |

The high infection/carriage rates reflect that more than one pathogen was recovered from many individuals in the study. We computed, based on observed recovery rates for each pathogen, the expected frequency of recovering combinations of DEC pathogens or *Salmonella*. Figure 4 shows that most co-infections were detected at the expected level but the observed of EAEC + EPEC, EAEC + ETEC and EAEC+ *Salmonella* co-infections exceeded expectation among controls, but not cases (p< 0.0025). Indeed, Salmonella were always recovered with a DEC isolate from controls and in one out of three cases, this was ETEC.

Table 3 shows the bacterial species isolated from samples during the study period. In addition to *E. coli*, some of which are pathogens, other explicit pathogens recovered were *Yersinia* and *Salmonella*. As for DEC, pathogenic genera *Salmonella* and *Yersinia* were not associated with diarrhea. However, they were proportionately more common in cases and controls and uncommonly recovered. There was a greater diversity of non-enteric pathogen genera in controls, compared to cases and *Klebsiella*, an abundant and commonly carried commensal, was detected significantly less commonly in cases than controls.

### Risk factors for DEC carriage

Age was not significantly associated with ETEC infection but ETEC was more commonly isolated from controls among younger children and cases in older age groups (Figure 3). Mother’s Education level was significantly associated with EAEC (p< 0.05) while the logistic regression shows that children with EAEC infection were more likely to have mothers with no education. The logistic regression of the Level of Mother’s Education shows that children whose mothers did not have secondary education were less likely to have EAEC infection (P<0.05, Supplementary Table 1). Bivarate analysis of parameters describing nutritional status of children and the household water sources of children from whom DEC pathotypes were recovered did not reveal any variables were associated with DEC recovery (Supplemental Table 2).

**Figure 3:**
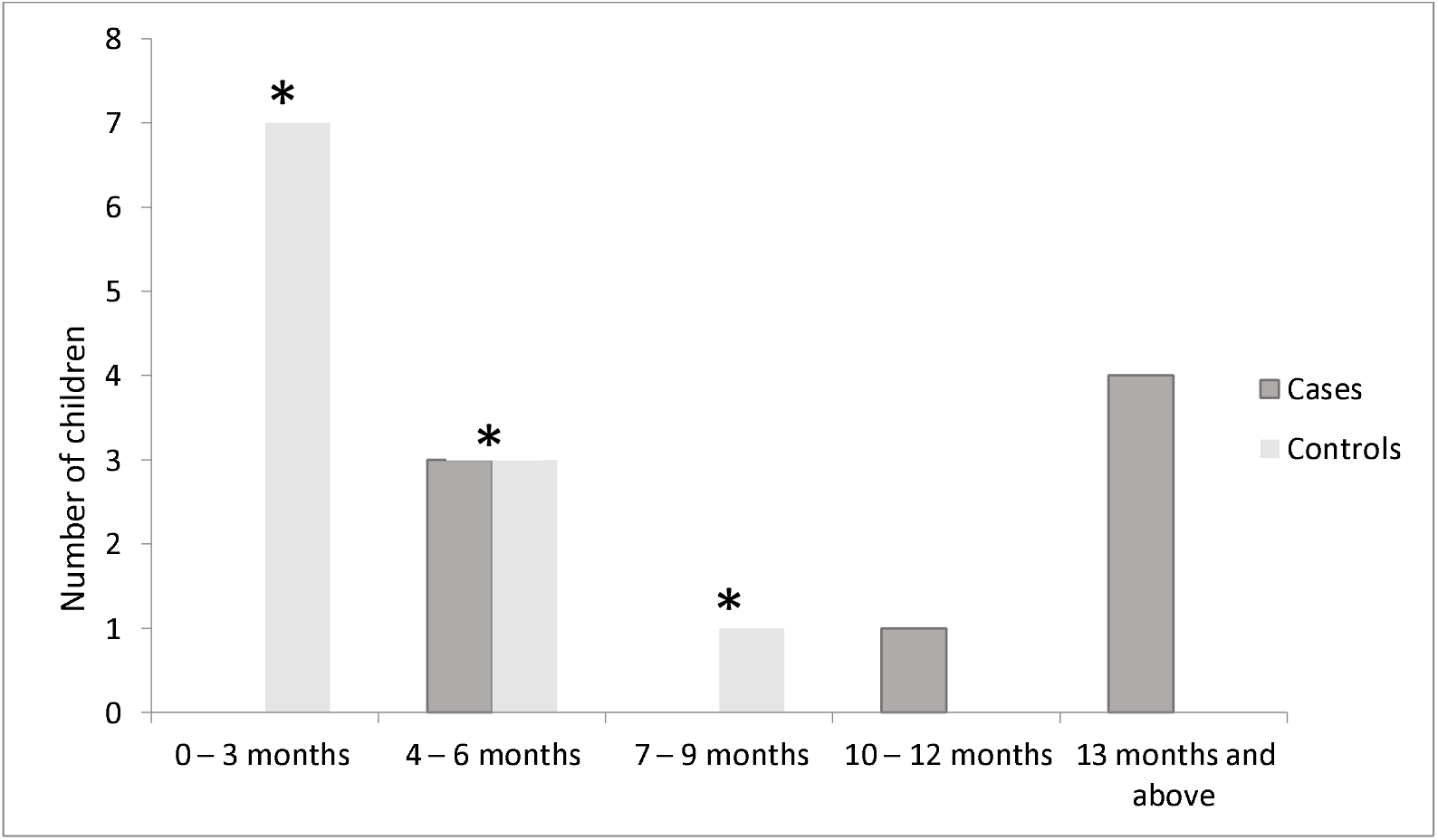
Age distribution children from whom ETEC was recovered (* indicates ETEC was recovered significantly more commonly from controls, compared with children with diarrhea p< 0.05).

**Figure 4:**
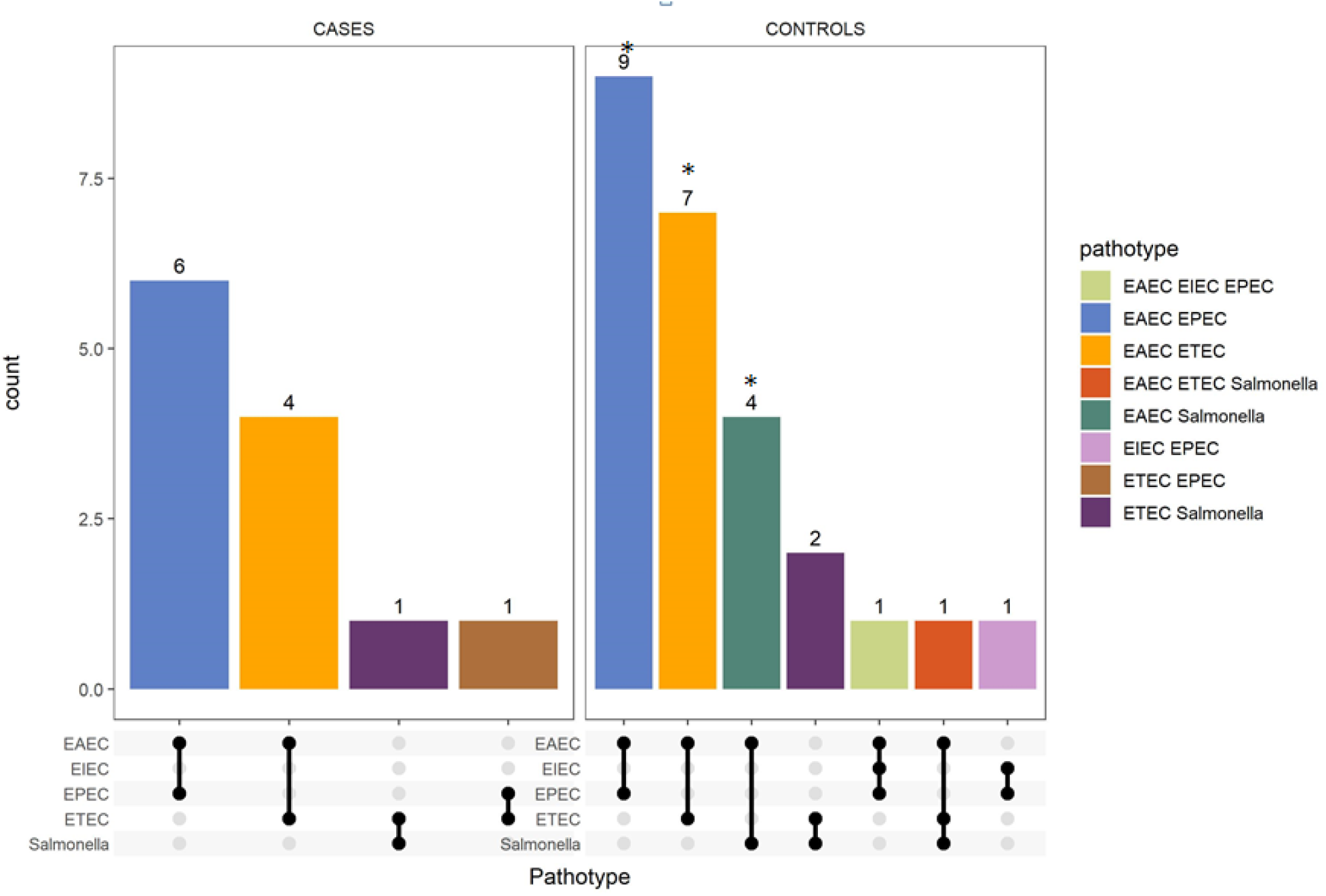
Number of patients co-infected with more than one DEC or *Salmonella enterica* pathotype (* indicates that the observed frequency of the co-infection exceeded expectation based on recovery of single infections among controls, but not cases, p ≤ 0.0025).

### EAEC carriage clusters

As Table 2 shows, cases were unevenly reported from the participating primary health care centers. The overall median number of cases recruited per month across the study was 3, but the range included months with up to nine enrollments. In months January-February 2016, September 2016, June 2018 and January-February (2019), the number of cases recruited was above two standard deviations of the mean of 3. Within these months EAEC isolation was over-represented and identical or near identical EAEC isolates were recovered from two individuals within each of these clusters (Table 2). However, at least one of the pairs in the cluster was a control, so that none of the detected clusters would be considered a disease outbreak *per se*. All the clusters occurred in drier months (rainfall < 4 mm).

**Table 2:** List of EAEC clusters.

| EAEC strains | No of individuals involved | Same or different household | *E. coli* ST | Study participants | Enrollment site (Primary care clinic) | Month/year of isolation |
| --- | --- | --- | --- | --- | --- | --- |
| LLH051B/LLH059E | 2 | Yes | 501 | Controls | Lalupon | Jan 2016, Feb 2016 |
| LWD015H / LLH054C | 2 | No | 648 | Case, Control | Lalupon | Jan 2016, Feb 2016 |
| MNH226F/MNH225J | 2 | Yes | 69 | Controls | Monotan | Jun 2018, Jun 2018 |
| MNH227C /MND75D | 2 | Yes | 40 | Case, Control | Monotan | Jun 2018, Jul 2018 |
| MNH224F/JKD76F | 2 | No | 1808 | Case, Control | Monotan, Ejioku | Jun 2018, Jun 2018 |
| LLH316ABCDE/LLH319C | 2 | Yes | 8746 | Controls | Lalupon | Feb 2019, Mar 2019 |
| LLH311A/LLH315C | 2 | Yes | 10 | Controls | Lalupon | Jan 2019, Feb 2019 |
| LLH315G/LLH311G | 2 | Yes | 1722 | Controls | Lalupon | Jan 2019, Feb 2019 |

### Total and coliform counts from sentinel water samples

Total counts of the water samples collected from 15 wells in the study area approached or exceeded 10^3^ cfu/mL and coliform counts ranged between 10^1^ and 10^4^ cfu/mL (Table 4). The presence of *E. coli*, indicating recent fecal contamination was recorded in 10 out of 14 of the wells. Salmonellae were enriched for but not cultured from any of the wells. As shown in table 4, a high total count was recorded from our control sample, UIH202, from a local water bottling plant, no coliforms or *E. coli* were detected.

**Table 3:** Enterobacterales and related genera isolated at least once from study cases and controls.

| Isolates | Cases<br>(n=120) | Controls<br>(n=357) | Total | P-value | Odds<br>ratio | CI (lower- Upper) |
| --- | --- | --- | --- | --- | --- | --- |
| Genus *Escherichia* | 106 | 307 | 413 | 0.6202 | 1.2331 | 0.6552-2.3210 |
| Genus *Acinetobacter* | 5 | 12 | 17 | 0.7759 | 1.2494 | 0.3375-3.9113 |
| Genus *Citrobacter* | 2 | 17 | 19 | 0.1795 | 0.3390 | 0.0375-1.4649 |
| Genus *Enterobacter* | 3 | 28 | 32 | 0.0516 | 0.3013 | 0.0577-1.0053 |
| Genus *Klebsiella* | 8 | 79 | 87 | 0.0001 | 0.2514 | 0.1176-0.5372 |
| Genus *Providencia* | 1 | 5 | 6 | 1.0000 | 0.5916 | 0.0124-5.3682 |
| Genus *Salmonella* | 3 | 6 | 9 | 0.5682 | 1.4986 | 0.2388-7.1492 |
| Genus *Serratia* | 3 | 24 | 27 | 0.1088 | 0.3564 | 0.0675-1.2063 |
| Genus *Yersinia* | 1 | 2 | 3 | 1.0000 | 1.4902 | 0.0251-28.8643 |
| Genus *Ewingella* | 1 | 0 | 1 | 0.2516 | -1.000 | -1.0000- -1.0000 |
| Genus *Hafnia* | 2 | 12 | 14 | 0.3789 | 0.4153 | 0.0930-1.8542 |
| Genus *Kluyvera* | 2 | 1 | 3 | 0.1574 | 6.0339 | 0.5422-67.1484 |
| Genus *Morganella* | 2 | 7 | 9 | 1.0000 | 0.8475 | 0.1736-4.1362 |
| Genus *Proteus* | 0 | 7 | 7 | 0.2001 | 0.0000 | -1.0000- -1.0000 |
| Genus *Stenotrophomonas* | 0 | 1 | 1 | 1.0000 | 0.0000 | -1.0000- -1.0000 |

**Table 4:**
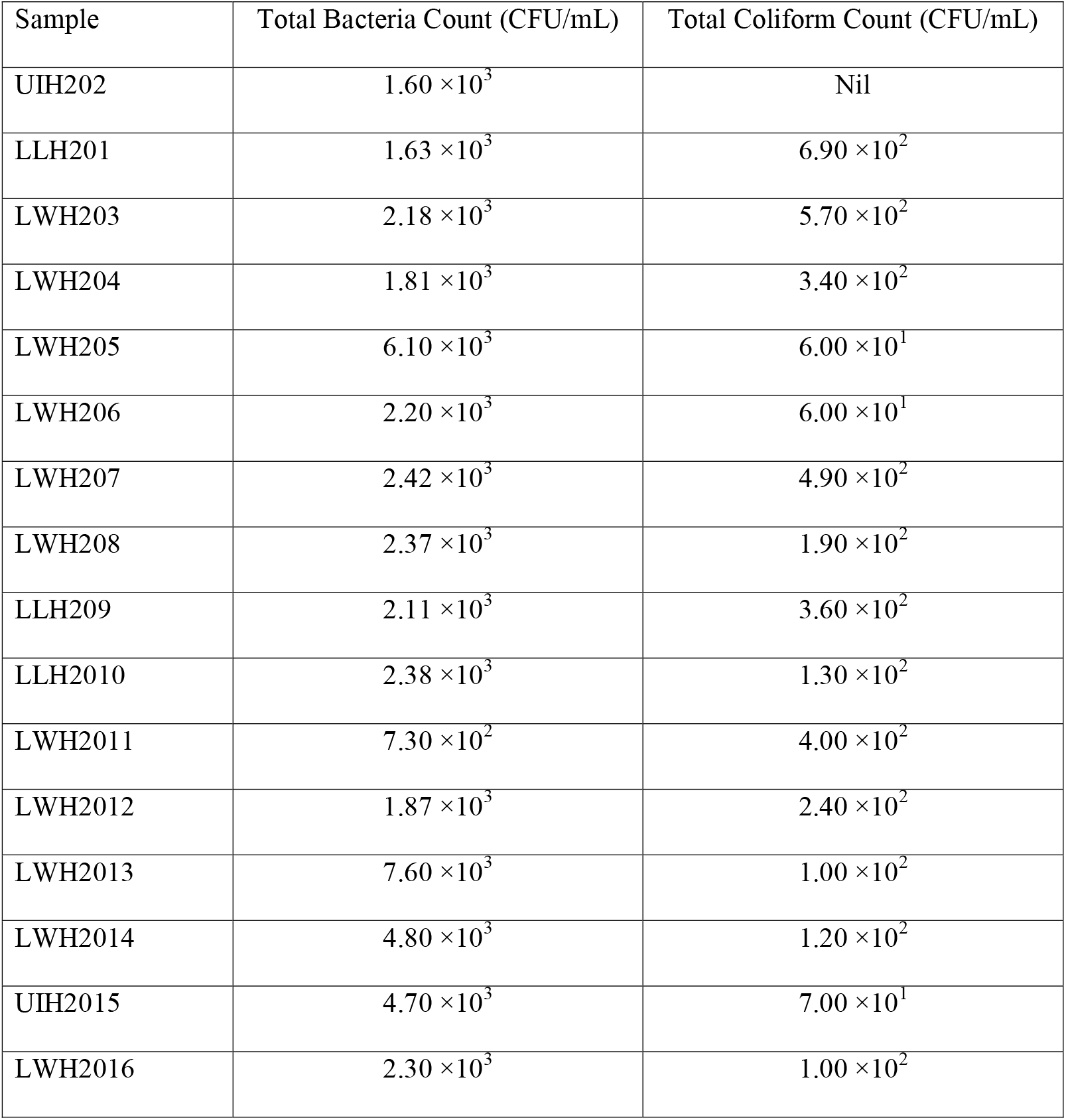
Total Coliform and total bacterial counts from sentinel water samples.

## Discussion

This study sought DEC pathotypes in specimens from 120 children that were taken to local primary health care centers with diarrhea and 357 controls. Our study participants were recruited from primary care centers and therefore illnesses largely represent the common bouts of mild diarrhea that contribute significantly to overall burden but are unlikely to reach hospitals where most diarrheal epidemiology studies are conducted.^7,13-17,25,41^ We isolated up to 10 colonies per specimen, ultimately recovering *E. coli* at least once from 106 (88.3%) in cases and 307 (86) controls. We additionally enriched for *Salmonella*. As is typical in studies of this nature, *E. coli* was the predominant enterobacterales species recovered. The diversity of enteric Gram negative bacteria was less in faecal specimens from children with diarrhea than controls with common enteric commensals such as *Klebsiella, Enterobacter* and *Citrobacter* spp. recovered less commonly from cases than controls. In addition to *E. coli*, some of which are pathogens, other unequivocal pathogens recovered were *Yersinia* (for which we did not enrich) and *Salmonella* (directly or after selenite broth enrichment). Although these genera are commonly associated with diarrhea in a range of studies, in this study they were proportionately more common in cases than controls but they were uncommonly recovered and not associated with diarrhea p > 1.1. This is reflective of the epidemiology we observed for DEC pathotypes as well and reflective of the high levels of enteric pathogen carriage we documented.

Salmonellae, Yersinae, EAEC, ETEC, EPEC and EIEC were isolated in this study but showed no association with disease, ^17,20,42-44^ (EHEC and STEC, which commonly do not feature in studies of under-five diarrhea in Africa, were not recovered). Screening multiple isolates per individual and the sensitivity of WGS for identifying *E. coli* subtypes, including atypical strains, allowed us to detect 537 DEC isolates in all, a much higher detection rate than in comparable studies and may also account for our high detection rate in controls. Ultimately, at least one DEC isolate was recovered from 77 (64.2%) cases and 220 (61.6%) controls.

A hybrid pathotype contains virulence or other loci that would lead to it being classified as more than one DEC pathotype, sometimes conferring greater virulence. Hybrids have been associated with outbreaks ^45-47^, however outbreak investigations typically prompt more comprehensive isolate characterization and would therefore be more likely to detect hybrids. In this study, 32 (2.6%) DEC isolates were hybrid DEC pathotypes. EAEC/EPEC hybrids, similar to an EAEC/ atypical-EPEC hybrid with the *eae* and *aagR* gene reported from Brazil in 2021^47^ were identified in 10 specimens (4 from cases and 6 from controls, NS). We additionally detected EAEC/ETEC in 16 specimens (5 cases and 7 controls, NS), ETEC/EPEC in 2 case specimens and EAEC/ EIEC in a control specimen.

Enteroaggregative *E. coli* (EAEC), which increasingly sought in diarrheal epidemiology studies and featuring prominently in those performed in Africa, ^7,17,21,25,31,48,49^ was the most commonly isolated DEC pathotype. EAEC is commonly associated with diarrhea in epidemiological studies in Africa,^7,17,31,41,^ but occasionally is not.^25,48,49^ High abundance of EAEC among both cases and controls has been seen previously in Nigeria,^17,18,21^ and was a key finding at multiple locations in the MAL-ED study in Brazil, Bangladesh, Tanzania India, Peru, South Africa, Pakistan, and Nepal.^50^ There is evidence that EAEC carriage may, in addition to or instead of diarrhea, lead to nutrient malabsorption,^7,50^ and therefore the high carriage rates we document are of concern, even in the absence of an association with frank diarrhea. Altogether EAEC was isolated from 281 (58.9%) children in this study including 95% of 296 children from whom DEC were recovered. EAEC was recovered with another DEC pathotype from 23 specimens and, in the case of EPEC, more commonly than combined probabilities would predict (p=0.0025). Chattaway et al (2013)^51^ have also reported that certain other pathogen combinations are common with EAEC and, while their significance is unclear, these combinations warrant further investigation.

The detection rate for ETEC in this study was high compared to previous studies in the region.^18^ ETEC were recovered from 8 (6.7%) of cases and 11 (3.1%) of controls (p= 0.0823, not significant). ETEC *sta* gene, encoding the heat stable enterotoxin, which is more commonly associated with diarrhea, ^7^ was however only detected in isolates from cases and one isolate harbored both the *ltcA* and *sta* genes. ETEC was more common in younger controls and older cases. Bivariate analysis confirmed age group associations with ETEC infection, showing association with diarrhea in children aged nine months and below (p<.0.01). Weaned children may lose protection from secretory IgA present in breastmilk, which has previously been reported to be protective for ETEC. ^55^ Increased exposure of older children may occur with weaning, and crawling, and consequent ingestion of infected material from food, drink and their surroundings may amplify their risk.^54^ However, our data do show that young children are exposed to ETEC and are simply not sickened by it. The poor quality of household water, which may be used for washing babies (discussed later) may be one of many unavoidable risk factors. In contrast to ETEC, the much higher carriage rates for EAEC and the age-inspecific risk, may in turn suggest that breastmilk protection does not occur for EAEC. It is however important to highlight that exclusive breastfeeding was not found to be protective against diarrhea or carriage of any of the pathotypes in this study. However non-exclusive breastfeeding was not queried.

EPEC are pathogens almost exclusively associated with childhood diarrhea. In the current study, EPEC was isolated from 7 (5.8%) case samples and 16 (4.5%) controls. Two control and one case isolate were typical EPEC with both *eae* (with other LEE genes) and *bfpA* genes. Atypical EPEC strains carrying LEE genes including *eae*, but lacking *bfpA* gene, were isolated from seven cases and 16 controls. EPEC have been historically prominent in Nigeria,^14,16^ based on reports from studies that used serotyping but more recent studies using hybridization,^17^ and PCR, ^18-20,56,57^ like our current WGS study have found this pathotype much less common suggesting that earlier results may be due to methodological artefacts.

*Shigella* has been reported as a principal etiological agent of diarrheal disease in African countries and has consequently been prioritized for vaccine development.^58-62^ EIEC, which have similar virulence factors may account for at least some of the global burden of *Shigella*.^17,41,53^ In the current study, only 1(0.3%) *Shigella sonnei* isolate (from a control) and no EIEC were recovered. This study also included only four cases of bloody diarrhea, two reported by patient care-givers, two others detected by fecal occult blood testing and none from which *Shigella* or EIEC were recovered. The relatively low recovery of *Shigella* in this study may be connected to our focus on largely mild diarrhea, treated close to home at primary health care facilities. Other similarly focused studies have made comparable findings.^17,63^ Reports in the literature largely focus on the moderate to severe diarrhea as likely to reach hospitals and *Shigella* may produce most disease in those patients.

A recent study, ^64^ in Cape Town, South Africa, found water sources and storage as a risk factor for diarrhea, likewise other African studies.^65-67^ We observed that household and shared wells were the most common water source used by residents in the areas served by enrollment primary helath care centers (Supplemental Table 3). When we sampled and analyzed water quality from 14 wells, we observed that all samples analyzed had high bacterial and coliform loads (exceeding the 20 CFU/mL and <1 CFU/mL respectively acceptable for potable water ^68^ and *E. coli* the standard indicator of faecal contamination, was isolated from 11 (78.6%) samples. Thus our findings demonstrate that improvements in water and sanitation should be a primary priority for intervention. Even when children are exclusively breast fed or fed water sources that are presumed to be improved (such as bottled or bagged water), household water is used for bathing and washing and could place very young children at risk.

In conclusion, this study found worrisomely high carriage rates for DEC and other bacterial pathogens with known etiologic roles in diarrhea. Children in northern Ibadan are commonly colonized with EAEC and other bacterial pathogens including, ETEC, EPEC, *Shigella, Salmonella* and *Yersinia*, as well as with DEC hybrids. While we did not find associations with diarrheal disease in the face of these high carriage rates, pathogens are frequently recovered from children reporting to primary health care centers with diarrhea. Asymptomatic colonization is common and becomes more pronounced after weaning. Moreover, identical EAEC strains were frequently recovered from children residing in different households suggesting that feco-orally transmitted pathogens are circulating. No well water sampled from a selection of households in the vicinity of the primary care centers was found to be potable and improving access to water and sanitation may be critical for protecting children from the less overt consequences of enteric pathogen carriage in northern Ibadan.

## Data Availability

the European Nucleotide Archive (ENA) under the project ID PRJEB8667 (https://www.ebi.ac.uk/ena/browser/view/PRJEB8667).

## ACKNOWLEDGEMENTS

The authors are grateful to David A. Kwasi, Anderson O. Oaikhena, Faith I. Oni, Taiwo Badejo, Anthony Underwood, Rotimi Dada, Ayorinde Afolayan, Erkison Odih, Catherine Ladipo and A Oladipo Aboderin for technical assistance and helpful comments. We thank Ola Aduroja and Chukwuemeka Nwimo for assistance with statistics and the staff of the Primary Health Centers where patients were recruited.

