## Supplementtal tables for "Epidemiology and risk factors for diarrhoeagenic *Escherichia coli* carriage among children in northern Ibadan, Nigeria"

**Supplementary Table 1: Social Demographic Characteristics of Children from whom DEC pathotypes were recovered**

|  |  | **EAEC** | | | | **ETEC** | | | | **EPEC** | | | |
| --- | --- | --- | --- | --- | --- | --- | --- | --- | --- | --- | --- | --- | --- |
| **Variables** |  | **Cases** | **Controls** | **X^2^** | **P-value** | **Cases** | **Controls** | **X^2^** | **P-value** | **Cases** | **Controls** | **X^2^** | **P-value** |
| Age | 0 – 3 months | 29 (61.7) | 148 (56.1) | 2.861 | 0.581 | 0 (0.0) | 7 (2.7) | 23.827 | **<0.01** | 3 (6.4) | 12 (4.5) | 4.156 | 0.385 |
|  | 4 – 6 months | 12 (48.0) | 37 (60.7) |  |  | 3 (12.0) | 3 (4.9) |  |  | 1 (4.0) | 3 (4.9) |  |  |
|  | 7 – 9 months | 11 (57.9) | 13 (59.1) |  |  | 0 (0.0) | 1 (4.5) |  |  | 3 (15.8) | 1 (4.5) |  |  |
|  | 10 – 12 months | 9 (75.0) | 4 (80.0) |  |  | 1 (8.3) | 0 (0.0) |  |  | 0 (0.0) | 0 (0.0) |  |  |
|  | 13 months and above | 11 (64.7) | 3 (60.0) |  |  | 4 (29.4) | 0 (0.0) |  |  | 0 (0.0) | 0 (0.0) |  |  |
| Sex | Male | 33 (57.9) | 118 (57.8) | 0.011 | 0.916 | 2 (5.3) | 5 (2.5) | 1.825 | 0.177 | 4 (7.0) | 10 (4.9) | 0.369 | 0.543 |
|  | Female | 39 (61.9) | 87 (56.9) |  |  | 6 (9.5) | 6 (3.9) |  |  | 3 (4.8) | 6 (3.9) |  |  |
| Mother’s School Education Level | No education | 4 (40.0) | 12 (44.4) | 7.812 | **0.05** | 2 (20.0) | 0 (0.0) | 0.380 | 0.944 | 0 (0.0) | 2 (7.4) | 1.837 | 0.607 |
|  | Primary | 7 (46.7) | 42 (58.3) |  |  | 2 (13.3) | 2 (2.8) |  |  | 3 (20.0) | 2 (2.8) |  |  |
|  | Secondary | 33 (56.9) | 99 (56.3) |  |  | 2 (5.2) | 7 (4.0) |  |  | 3 (5.2) | 10 (5.7) |  |  |
|  | Tertiary | 28 (75.7) | 52 (63.4) |  |  | 2 (5.4) | 2 (2.4) |  |  | 1 (2.7) | 2 (2.4) |  |  |
| Location | Alakia | 3 (50.0) | 7 (50.0) | 2.276 | 0.685 | 0 (0.0) | 0 (0.0) | 8.933 | 0.063 | 0 (0.0) | 0 (0.0) | 3.114 | 0.539 |
|  | Ejioku | 1 (50.0) | 4 (40.0) |  |  | 0 (0.0) | 2 (20.0) |  |  | 0 (0.0) | 0 (0.0) |  |  |
|  | Elewi-odo | 15 (62.5) | 6 (60.0) |  |  | 1 (8.3) | 0 (0.0) |  |  | 1 (4.2) | 1 (10.0) |  |  |
|  | Lalupon | 31 (59.6) | 149 (57.8) |  |  | 3 (5.8) | 3 (4.6) |  |  | 2 (5.6) | 14 (5.4) |  |  |
|  | Monatan | 22 (61.1) | 39 (60.0) |  |  | 4 (11.1) | 6 (2.3) |  |  | 4 (7.7) | 1 (1.5) |  |  |

*Empty Cells are either references or cells with zero values

**Supplementary Table 2: Association of exclusive breastfeeding and nutritional status of children with DEC pathotypes**

|  |  | **EAEC** | | | | **ETEC** | | | | **EPEC** | | | |
| --- | --- | --- | --- | --- | --- | --- | --- | --- | --- | --- | --- | --- | --- |
| **Variables** |  | **Cases** | **Controls** | **X^2^** | **P-value** | **Cases** | **Controls** | **X^2^** | **P-value** | **Cases** | **Controls** | **X^2^** | **P-value** |
| BMI | Underweight (<18.5) | 56 (64.4) | 157 (57.7) | 2.186 | 0.535 | 7(9.2) | 10 (3.7) | 2.489 | 0.715 | 7 (8.0) | 10 (3.7) | 2.344 | 0.504 |
|  | Normal (18.5 – 24.9) | 15 (51.7) | 41 (57.7) |  |  | 1 (3.4) | 1 (1.4) |  |  | 0 (0.0) | 5 (7.0) |  |  |
|  | Overweight (25.0 – 29.9) | 1 (50.0) | 1 (25.0) |  |  | 0 (0.0) | 0 (0.0) |  |  | 0 (0.0) | 1 (25.0) |  |  |
|  | Obese (30.0 and above) | 0 (0.0) | 5 (62.5) |  |  | 8 (7.5) | 0 (0.0) |  |  | 0 (0.0) | 0 (0.0) |  |  |
| Exclusive breastfeeding | Yes | 34 (63.0) | 142 (55.7) | 0.447 | 0.504 | 4(9.3) | 9 (3.5) | 0.249 | 0.618 | 2 (3.7) | 10 (3.9) | 1.683 | 0.195 |
|  | No | 38 (57.6) | 63 (61.8) |  |  | 4 (6.1) | 2 (2.0) |  |  | 5 (7.6) | 6 (5.9) |  |  |
| **Height for Age** | Severely Stunted | 16 (59.3) | 49 (51.6) | 4.251 | 0.373 | 3 (11.1) | 4 (4.2) | 3.183 | 0.528 | 1 (3.7) | 3 (3.2) | 4.114 |  |
|  | Stunted | 9 (45.0) | 32 (61.5) |  |  | 2 (15.0) | 1 (1.9) |  |  | 0 (0.0) | 2 (3.8) |  |  |
|  | Tall | 43 (63.2) | 106 (57.6) |  |  | 3 (4.4) | 4 (2.2) |  |  | 5 (7.4) | 9 (4.9) |  |  |
|  | Very Tall | 4 (80.0) | 16 (72.7) |  |  | 0 (0.0) | 2 (9.1) |  |  | 1 (20.0) | 2 (9.1) |  |  |
| **MUAC** | Healthy | 50 (59.5) | 62 (53.9) | 2.096 | 0.718 | 8 (10.7) | 4 (3.5) | 7.933 | 0.094 | 4 (4.8) | 7 (6.1) | 7.214 |  |
|  | Malnourish | 2 (66.7) | 4 (57.21) |  |  | 0 (0.0) | 1 (14.3) |  |  | 0 (0.0) | 0 (0.0) |  |  |
|  | Not applicable | 17 (58.6) | 136 (59.4) |  |  | 0 (0.0) | 5 (2.2) |  |  | 2 (6.9) | 8 (3.5) |  |  |
|  | Severely Malnourished | 3 (75.0) | 3 (60.0) |  |  | 0 (0.0) | 1 (20.0) |  |  | 1 (25.0) | 1 (20.0) |  |  |
| **Weight for Age** | Healthy | 40 (54.1) | 138 (58.0) | 8.720 | 0.068 | 5 (8.1) | 3 (1.3) | 6.008 | 0.199 | 3 (4.1) | 9 (3.8) | 2.772 |  |
|  | Overweight | 2 (50.0) | 0 (0.0) |  |  | 0 (0.0) | 0 (0.0) |  |  | 0 (0.0) | 0 (0.0) |  |  |
|  | Severely | 18 (69.2) | 27 (50.0) |  |  | 3 (11.5) | 4 (7.4) |  |  | 3 (11.5) | 3 (5.6) |  |  |
|  | Underweight | 12 (75.0) | 40 (65.6) |  |  | 0 (0.0) | 4 (6.6) |  |  | 1 (6.3) | 4 (6.6) |  |  |

*Empty Cells are either references or cells with zero values

**Supplementary Table 3: Water Sources of Children with DEC pathotypes**

|  |  | **EAEC** | | | | **ETEC** | | | | **EPEC** | | | |
| --- | --- | --- | --- | --- | --- | --- | --- | --- | --- | --- | --- | --- | --- |
| **Variables** |  | **Cases** | **Controls** | **X^2^** | **P-value** | **Cases** | **Controls** | **X^2^** | **P-value** | **Cases** | **Controls** | **X^2^** | **P-value** |
| Source of drinking water in the household | Bottled water | 13 (61.9) | 27 (54.0) | 4.491 | 0.610 | 1 (4.8) | 3 (6.0) | 2.857 | 0.827 | 1 (4.8) | 4 (8.0) | 6.318 | 0.388 |
|  | Well | 16 (41.0) | 104 (59.8) |  |  | 2 (5.1) | 8 (4.6) |  |  | 2 (5.1) | 7 (4.0) |  |  |
|  | Bagged water | 17 (70.8) | 16 (47.1) |  |  | 2(12.5) | 0 (0.0) |  |  | 1 (4.2) | 2 (5.9) |  |  |
|  | Borehole | 4 (66.7) | 15 (53.6) |  |  | 0 (0.0) | 0 (0.0) |  |  | 2 (33.3) | 2 (7.1) |  |  |
|  | Boiled water | 9 (60.0) | 21 (60.0) |  |  | 1 (6.7) | 0 (0.0) |  |  | 1 (6.7) | 0 (0.0) |  |  |
|  | Tap water | 12 (85.7) | 22 (64.7) |  |  | 2 (14.3) | 0 (0.0) |  |  | 0 (0.0) | 1 (2.9) |  |  |
|  | River or stream water | 1 (100.0) | 0 (0.0) |  |  | 0 (0.0) | 0 (0.0) |  |  | 0 (0.0) | 0 (0.0) |  |  |
| Source of household water for general use | Well | 16 (41.0) | 104 (59.8) | 4.491 | 0.610 | 2 (5.1) | 8 (4.6) | 2.857 | 0.827 | 2 (5.1) | 7 (4.0) | 6.318 | 0.388 |
|  | River or stream | 1 (100.0) | 0 (0.0) |  |  | 0 (0.0) | 0 (0.0) |  |  | 0 (0.0) | 0 (0.0) |  |  |
|  | Borehole | 4 (66.7) | 15 (53.6) |  |  | 0 (0.0) | 0 (0.0) |  |  | 2 (33.3) | 2 (7.1) |  |  |
|  | Tap water | 12 (85.7) | 22 (64.7) |  |  | 2 (14.3) | 0 (0.0) |  |  | 0 (0.0) | 1 (2.9) |  |  |
|  | Boiled water | 9 (60.0) | 21 (60.0) |  |  | 1 (6.7) | 0 (0.0) |  |  | 1 (6.7) | 0 (0.0) |  |  |
|  | Bagged water | 17 (70.8) | 16 (47.1) |  |  | 2 (12.5) | 0 (0.0) |  |  | 1 (4.2) | 2 (5.9) |  |  |

*Empty Cells are either references or cells with zero values
